# Development of a Suicide Prevention Life Gatekeeper Training Program in China: A Delphi study

**DOI:** 10.1101/2023.01.01.22284096

**Authors:** Chengxi Cai, Chen Yin, Yongsheng Tong, Diyang Qu, Yunzhi Ding, Daixi Ren, Peiyu Chen, Yi Yin, Jing An, Runsen Chen

**Author notes:** Contributed equally (Chengxi Cai, Chen Yin). Correspondence to: Dr. Runsen Chen or Dr. Jing An.

## Abstract

**Introduction:** Youth suicide has been a pressing public mental health concern in China, yet there is a lack of localised gatekeeper intervention programs developed for Chinese schools. Life Gatekeeper was the first systematically developed gatekeeper program which aimed to equip teachers and parents with knowledge, skills, and ability to identify and intervene students at high risk of suicide. This study aimed to achieve expert consensus on the content of this program.

**Methods:** The Delphi method was used to elicit consensus on statements essential to include in the training program. In the form of scoring existing statements and responding to open-ended questions, experts have the opportunity to evaluate the content, delivery form, feasibility, and overall feeling of the initial training program. Two Delphi rounds were conducted among Chinese panel members with diversified professional backgrounds in suicide research and practice. Statements were accepted for inclusion in the adjusted training program if they were endorsed by at least 80% of the panel.

**Results:** Consensus was achieved on 201 statements out of 207 statements for inclusion in the adapted guidelines for the gatekeeper programme, with 151 from the original questionnaire, and 50 generated from the comments of the panel members. These endorsed statements were used to develop the Life Gatekeeper training program.

**Conclusion:** This Delphi study provided an evidence base in developing of the first gatekeeper training program in China. We hope that the current study could pave the way for more evidence based suicide prevention programs in China. Further study is warranted to evaluate the effectiveness of the Life Gatekeeper training program.

## Introduction

Suicide has been a pressing global public health concern and a leading cause of death of young people aged between 10 to 24 years old (WHO, 2014; CDC, 2020), which imposes substantial economic burden on countries across the globe (Erskine et al., 2015; Kinchin & Doran, 2018; Lingeswaran, 2012; Ortiz-Prado et al., 2017). Although the overall suicide rate in China has declined significantly in the past few decades due to economic development and improvement in living standards (Cai et al., 2021), the proportion of adolescents at risk of suicide remains concerning. For example, a recent cohort study found that among student aged between 12 to 18 years old, the lifetime prevalence rate of suicidal thoughts ranged from 17.6% to 23.5%, whilst the prevalence rate for suicide plan ranged from 8.9% to 10.7%, and the prevalence rate of lifetime suicide attempts ranged from 3.4% to 4.6% (Liu et al., 2019). Similarly, another study of 12,733 Chinese students aged 9 to 18 years old found that 38.1% of female and 30.0% of male students reported suicide ideation (Tan et al., 2018). The Ministry of Education of the People’s Republic of China (2020, 2021) has recognised the need of promoting mental health services to reduce suicidal risks among Chinese students, and advocated a strong collaborative partnership between school and family, as well as between school and mental health services, to improve the ability to intervene with psychological crisis among students.

Adolescent suicide has detrimental impact not only on their parents and families, but also on their teachers and peers at school (Kolves et al., 2017; Kourkouta et al., 2019; Swanson & Colman, 2013). Meanwhile, these people are also referred to as ‘gatekeepers’ who have primary contact with at-risk students, who have the potential to identify warning signs, and hence intervene with the student before suicide took place (Isaac et al., 2009). Thus, school-based gatekeeper training program has been a widely used suicide prevention strategy, which aims to equip teachers, school personnel, parents and peers with the skills to recognise warning signs in students at risk of suicide, to communicate with the latter effectively, and to refer them to seek formal support (Cross et al., 2011; McKay et al., 2022; Totura et al., 2019). There have been a range of gatekeeper programs developed internationally, such as Question, Persuade, Refer (QPR; Quinnett, 1995; Wyman et al., 2008; Aldrich, Wilde & Miller, 2018), and Applied Suicide Intervention Skills Training (ASIST; Gould et al., 2013; Shannonhouse et al., 2017). Key components of such programs include psycho-education on myths and facts about suicide, warning signs, reducing stigma, and promoting gatekeeper behaviour of communicating and referring at risk students to seek professional psychological support. Overall, current gatekeeper programs have been found to reduce stigma, increase gatekeepers’ knowledge of suicide and self-efficacy to intervene (Holmes et al., 2021; Mo, Ko & Xin, 2018). However, the efficacy of gatekeeper programs in reducing suicide behaviour and in increasing gatekeeper behaviour remains unclear and lacks conclusive evidence (Kutcher, Wei & Behzadi, 2017; Torok et al., 2019). A recent meta analysis found that low quality of empirical findings for gatekeeper training could be contributed to small sample size and short follow-up time (Pistone et al., 2019). Moreover, Burnette, Ramchand and Ayer (2015) noted a largely unstudied gap between knowledge, beliefs, and skills learned in gatekeeper training, and their translation to actual gatekeeper behaviour. Recently, an emerging group of researchers have found that the theory of planned behaviour (TPB; Ajzen, 1991) could be used to predict intention (Aldrich, 2015; Servaty-Seib et al., 2013) and actual gatekeeper behaviour (Kuhlman et al., 2017; Kuhlman et al., 2021), thus the theoretical framework should be applied when developing an effective gatekeeper training program (Cox, 2021; Shemanski Aldrich & Cerel, 2009).

In summary, an evidence-based and effective suicide prevention is urgently needed in response to the pressing public health concern of youth suicide in China, yet there is a strikingly paucity for such research. To the best of our current knowledge, although there have been research on effectiveness of gatekeeper training among university students in China (Liang, Zhang & Xu, 2016; Zhao,Jing,Wang, 2010), no such study has been conducted among secondary or high school teachers or parents. There were studies about the development of a gatekeeper training program for school teachers and for university students and staff, yet these studies were mostly descriptive and did not present any empirical findings (Xu, Zhao & Fu, 2015; Xu & Xu, 2019). To the best of our knowledge, no study on gatekeeper training for parents in China could be found. Meanwhile, gatekeeper training programs developed in Western countries might not be appropriate to be implemented in Chinese schools directly without careful consideration of relevant sociocultural factors. For example, children born in rural areas might become left-behind children (LBC) if either one or both of their parents migrate to urban areas to earn a living (Su et al., 2013), and studies have found that parental migration has a detrimental impact on suicide behaviour of LBC (Fellmeth et al., 2018; Qu et al., 2021; Xiao et al., 2019), whilst the prevalence of suicide ideation and suicide attempt was found to be significantly higher among LBC compared to non-left-behind children (NLBC; Zhou et al., 2022; Chang et al., 2017). Futhermore, it was found that only 35% of Chinese schools in Beijing have qualified mental health counsellors, 20% school did not provide any psychological service, and the ratio between psychological service personnel and students was 1:1360 (Wang et al., 2015). The severe shortage of qualified counsellors, and other challenges such as lack of standardised training or evaluation of them means that it might not always be feasible to refer at-risk students to school counsellors as taught in existing gatekeeper training programs (Shi, 2018).

Given the lack of rigorously designed and localised gatekeeper training programs in China, especially with the need to take relevant ethical issues, sociocultural factors and implementation difficulties into consideration, the current study utilised the Delphi method as it could provide an evidence to develop such intervention by ‘systematically tapping’ into the expertise of a group of Chinese researchers and clinicians recognised in the field of suicide intervention (Minas & Jorm, 2010). Essentially, the aim of the Delphi method is to achieve consensus among experts on the proposed content, by an iterative process of analysing feedback from the experts and revising the proposed content (Diamond et al., 2014). Previously, this expert consensus method has been used in developing mental health first aid guidelines for suicide in China (Lu et al., 2020), and in developing other mental health interventions (Sharpe et al., 2020; Spain & Happé, 2020). Thus, this study aimed to use the Delphi method as an essential part of developing the first evidence-based, localised suicide gatekeeper training in China.

## Materials and Methods

### The Delphi method

The Delphi technique involves a group of experts making independent ratings of agreement with a series of statements through an iterative, multi-stage process. This systematic approach draws on the expertise of people working in specific areas and is applicable to provide guidance in a particular context. Delphi studies have commonly been used for the content development of mental health training programs, including culturally appropriate mental health first aid (Chalmers et al., 2014), suicide postvention guidelines for secondary schools (Cox et al., 2016), and Dos and Don’ts in Designing School-Based Awareness Programs for Suicide Prevention (Grosselli, 2021).

Delphi process was conducted to identify the core components of the school-based youth suicide prevention gatekeeper training program. Panel members were invited to review and rate their agreement with a range of initial statements and suggest any additional items that were not included in the original questionnaire. New statements suggested by panel members in Round 1 were included in Round 2 for all members to rate. After obtaining the data from Round 1, panel members received feedback on their responses and a summary of the overall ratings. Subsequently, they were able to re-rate in Round 2 where they had the opportunity to change or maintain their original rating in light of this feedback. First round of questionnaires took place between September and October, 2022, whilst the second round was conducted between October and November, 2022. The list of statements with substantial consensus in ratings generated from this process was included as the contents for the training program, and statements with low or conflicting ratings were discarded. This study was approved by The Institution review board of Tsinghua University (project number 20220128).

### Literature review

This search was used to identify information about the main contents of the training program, and search keywords were determined based on previous study (Mo et al., 2018). The keywords of ‘school-based’, ‘curriculum based’, ‘suicide prevention’, ‘suicide education’, ‘gatekeeper’, ‘teacher’, ‘staff’, ‘parent’ and their various synonyms were investigated in the databases of Google Scholar, Web of Science, PubMed and Chinese National Knowledge Infrastructure (CNKI).

### Questionnaire development and adaptation

The content of the intervention was initially developed based on reviewing existing literature relevant to school-based gatekeeper training for suicide prevention, followed up by discussion with school counsellors and experts in suicide prevention for practical insights in intervention development before the Delphi study.

Statements for the contents of the program were organized into eight common sections: (1) The severity of suicide among adolescents and the common feelings of suicidal persons, (2) Establish a correct understanding of suicide, (3a) Risk factors associated with suicide, (3b) Identify the warning signs of suicide, (4) The correct way to communicate suicide risk,(5) Assess suicide risk, (6) Make a safety plan, (7a - For Teacher’s Training Only) Teachers communicate with parents about their children’s suicide risk and find help for them, (7b - For Parent training only) Parents express support to their children and find resources for help, (8) In addition to the previously mentioned stigma and morbidity, other reasons that prevent children from seeking help or teachers or parents from providing help. First six modules of the program match key components of existing gatekeeper programs, and the latter two sections were developed as localised and innovative features of the Life Gatekeeper program. The teacher-specific and parent-specific modules were developed in response to the call for family-school partnership in suicide prevention, and the final module which encourages trainees to discuss potential barriers was in line with the theory of planned behaviour (Ajzen, 1991; Kuhlman et al., 2021), and with the aim to promote positive attitude and increased perceived behavioural control about performing gatekeeper behaviour.

After that, the research team members set up a working group, including experts in the field of mental health intervention project development and suicide prevention. The Working Group met regularly to discuss each possible statement extracted from the preliminary contents that may be applicable to this training program. We revised the statements to ensure that they could be understood by teachers and parents who lacked background knowledge of suicide prevention and were suitable for implementation. After several rounds of discussion and modification, the statements applicable to this intervention program constituted the initial Delphi study questionnaire.

### Panel formation

The chosen panel members have relevant suicide prevention or intervention professional experience. The experts are individually invited if they met any the following inclusion criteria.

a. The members of the Crisis Intervention Committee of the Chinese Association for Mental Health
b. Psychiatrists/psychotherapists working in medical institutions for more than five years, with clinical experience in suicide intervention.
c. Professors engaged in teaching and research of psychology (psychological crisis intervention) in colleges and universities.
d. School counselors who often participated in students’ psychological counseling and involved in suicide crisis management.
e. Crisis line operators with more than five years’ working experience in answering and managing the crisis intervention hotline.

### Data collection and analysis

At the beginning of each round of the Delphi study, an online link to participate in the survey was sent to all of the experts on the list. One week later, the experts who had not completed the questionnaire received an email to remind them. Each round of the study lasted for two weeks, and responses that exceeded the time limit were not obtained or included in the data analysis.

Panel members completed two rounds of questionnaires using the web-based survey platform “Wenjuanxing”. They were advised that their participation in the Delphi study would lead to the development of a gatekeeper training program for suicide prevention that was culturally, and contextually appropriate for teachers and parents in China. During the Round 1 and Round 2 questionnaires, panel members were instructed to rate each statement according to its importance for inclusion in the program, as well as feasibility for the training forms or achieving the training objectives, and appropriateness of the training materials. A five-point Likert scale was used for rating of above criteria, which included the options: importance (1□=□Least important, 2□=□unimportant, 3□= unsure/depends, 4□=□important, and 5□=□essential), feasibility(1□=□not feasible at all, 2□=□not feasible, 3□=□unsure/depends, 4□=□feasible and 5□=□completely feasible), and appropriateness(1□=□not appropriate at all, 2□=□not appropriate, 3□=□unsure/depends, 4□=□appropriate, and 5□=□completely appropriate).

After each round, responses were analyzed to calculate the percentage of the panel who rated an item as 4 or 5. According to previous similar studies (De Silva et al., 2016; Lu et al., 2020), the criteria for consensus was defined as 80% or more of the panels scoring an item as necessary (≥ 4). Statements that were endorsed by 80% or more of the panel members were included in the training guide immediately. Statements rated by 70-79% of the panel members as necessary were re-evaluated in the following round. Statements that were rated by less than 70% of the panel members as necessary were immediately excluded.

Following Round 1, all panel members were provided with a summary report that included a comparison of their own ratings against the overall response for each item. In the Round 2 questionnaire, panel members were asked to re-rate statements which were endorsed by 70-79% of panel members, and rate new statements created from open-ended questions from Round 1, to include in the training guide.

## Results

### Expert panel information

In Round 1, 34 of the 40 invited potential expert panel members agreed to participate in the study and completed the survey. 31 of them participated in Round 2 (retention rate□=□91%). All penal members were currently working in China and were from 13 provincial regions, including Beijing (35%), and Hubei (15%), Hunan, Shanxi (Central provinces), Jilin, Liaoning, Inner Mongolia, and Tianjin (Northern provinces), Shanghai, Zhejiang, and Shandong (Eastern provinces), Fujian and Guangdong (Southern provinces). The composition of participants represented various professional backgrounds. Most of them were school or university teachers (38%), psychiatrists (35%), psychotherapists (27%), psychologists (33%), academics (14%), and hotline operators (18%) (See Table 1 for more details).

**Table 1.**
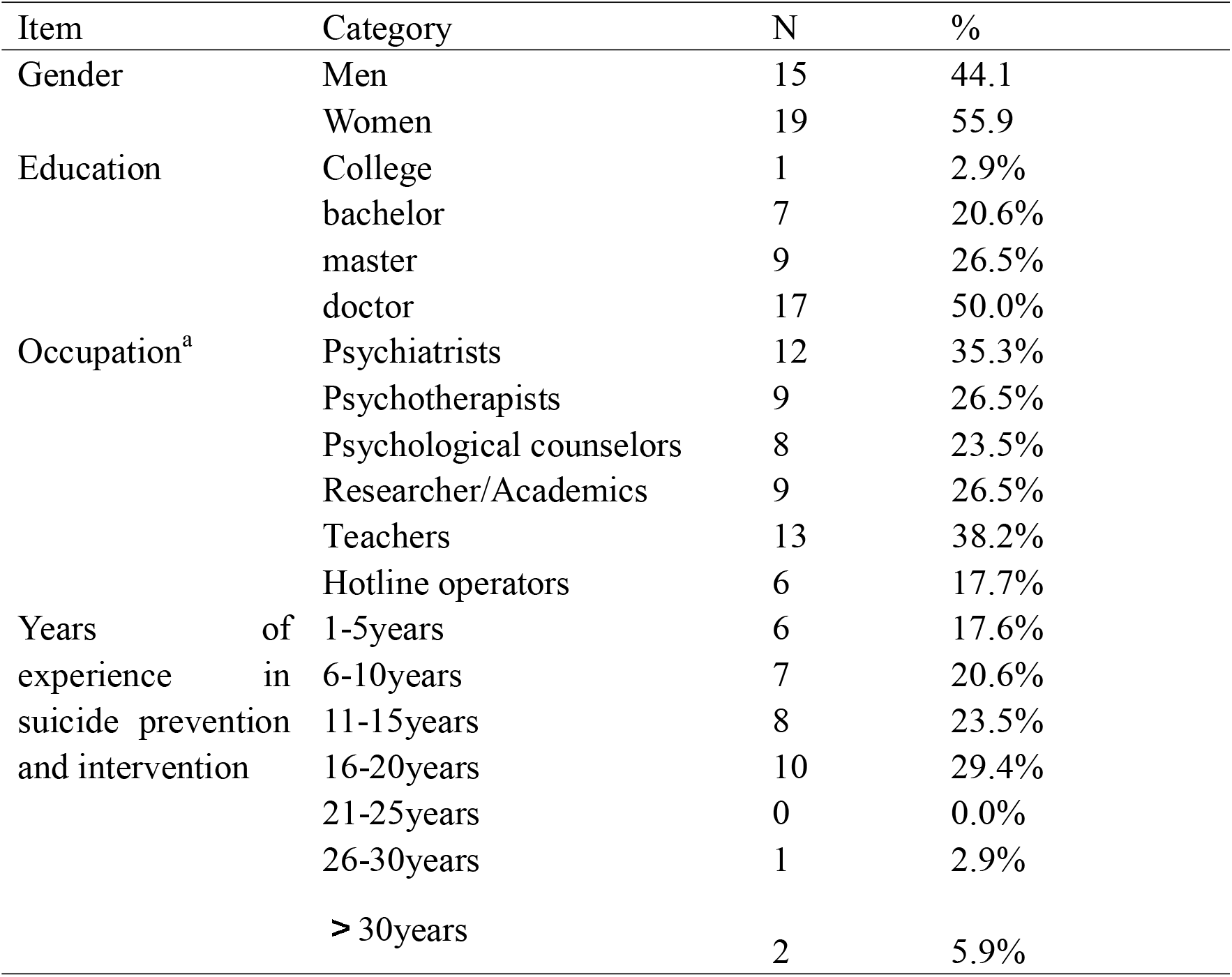
Characteristics of participants (n=34) a.including 13 participants with multiple occupations

## Endorsed statements

Figure 1 shows the number of statements to be endorsed, re-rated, rejected, and newly added in each Delphi round. Of the 157 items included in the first round, 151 were endorsed, 1 was excluded, and 5 needed to be re-rated. All of the 50 new statements developed from panel members’ comments collected through the Round 1 survey were endorsed in the Round 2 rating process. Of the 55 statements included in Round 2, all of the 5 re-rating statements were rejected and excluded. After the two survey rounds, 201 of the 207 statements assessed by the expert panel, were endorsed for inclusion in the gatekeeper training program.

**Figure 1.**
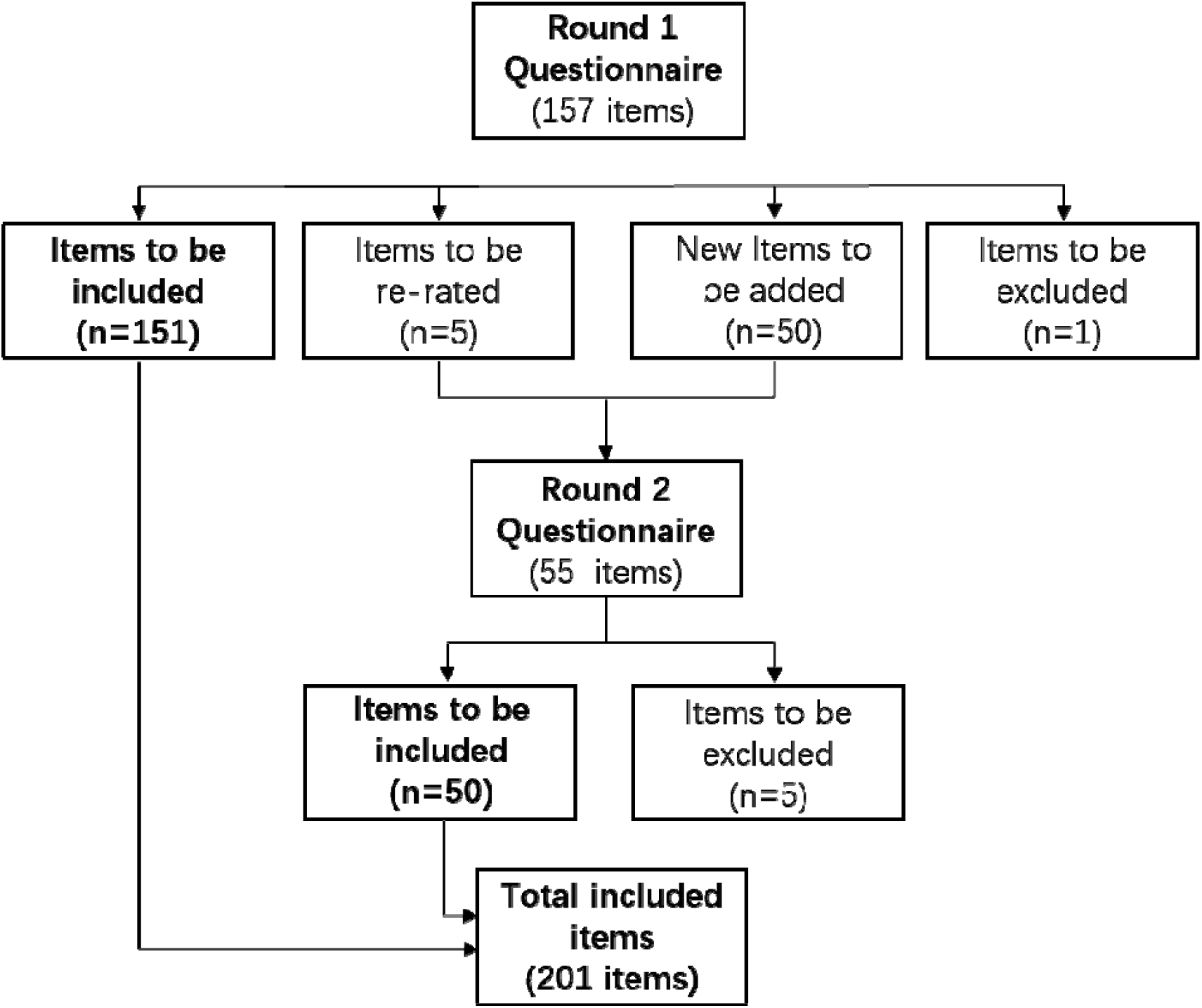
Flow of the delphi consensus process

The following are some examples of the new statements generated according to the open-ended text comments after each section, see Table 2. Table 3 shows examples of statements endorsed for inclusion in the training program.

**Table 2.**
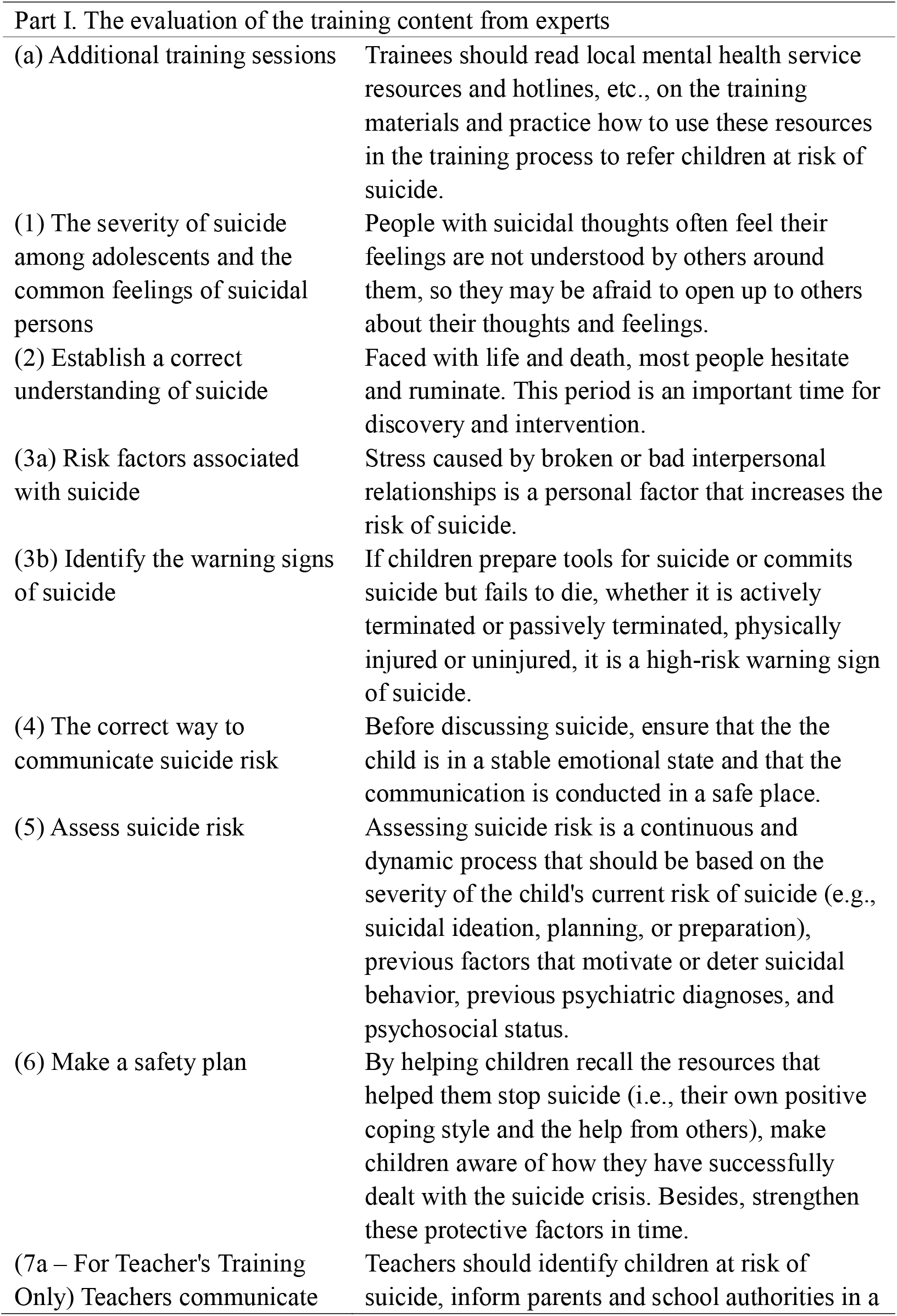
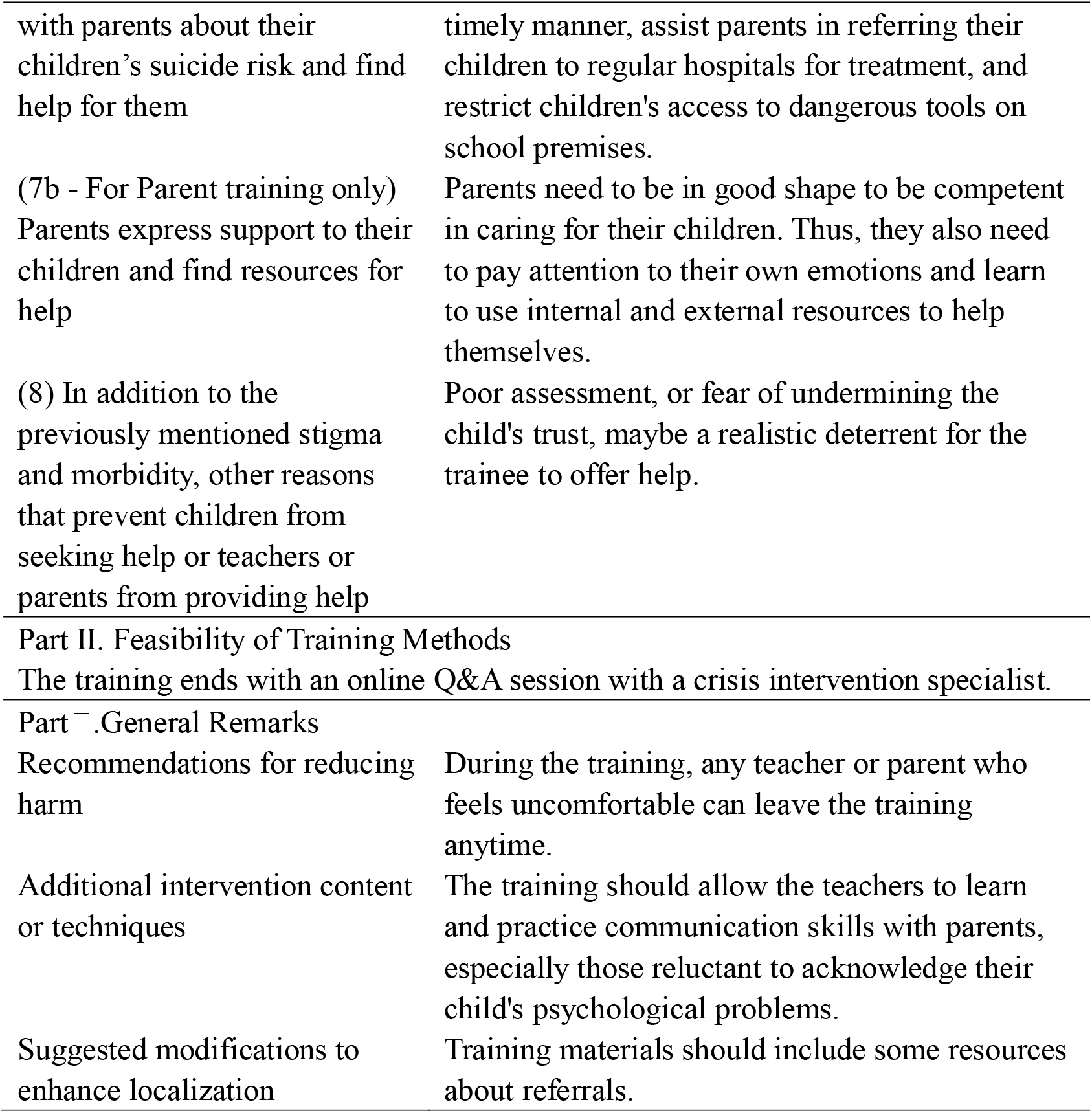
Example of the new items generated from suggestions and were endorsed by the panel

**Table 3.** Example of endorsed items and their agreement percentages.

| Part I. The evaluation of the training content from experts | Ratings (%) |
| --- | --- |
| (a) The importance of each chapter |  |
| Help trainees identify children at risk of suicide as early as possible by explaining suicide-related risk factors and early warning signs. | 100.0 |
| Trainees will practice how to talk about suicide and get timely feedback from companions during role play, which will help them feel more confident when talking about suicide in the future. | 100.0 |
| By practicing asking about suicidal thoughts, plans, tools, and methods, trainees will learn how to assess children's risk of suicide. | 100.0 |
| Part II. Feasibility of Training Methods |  |
| Watching videos (teaching-related knowledge through animation and video clips) | 91.2 |
| Part III. Feasibility of achieving the training objectives |  |
| Statistics, common misconceptions, risk factors, and warning signs related to suicide are presented in the form of videos. This is to help trainees deepen their understanding and memory of the relevant information. | 97.1 |
| Part IV. Suitability of training materials |  |
| This intervention will provide standardized training materials (i.e., intervention videos, manuals, appendices, training presentations, and materials for children and parents) to facilitate the generalization and ongoing learning of this intervention. | 100.0 |
| Part V. General Remarks |  |
| When training parents, parents from different backgrounds should be surveyed about their perceptions of suicide prevention in schools and their willingness or barriers to participating in the training. | 96.8 |

According to the results of two-rounds of the Delphi questionnaire, we have revised the content of the training program. Among them, Section I is used for the content development of the training project based on the importance of each section and specific item for inclusion in the program. Section II was used to adjust the form of the training, and the third part was used to confirm the experts’ evaluation of whether the existing training methods could achieve the desired objectives. The fourth part was used to modify the form and content framework of the training materials. In the fifth part at the end of the questionnaire, participants were asked to indicate their overall feelings about the training program, and whether they considered the program to be culturally suitable for the local context, or harmful. According to their comments, new items were generated to modify the existing training form and content, as well as the overall prerequisites and ways of providing the training program.

## Discussion

This is the first Delphi study conducted as part of the development of a school-based gatekeeper program in China. Overall, expert consensus achieved in the current study provided an evidence basis for the systematic development of the Life Gatekeeper program, which is of great significance given the current lack of such school-based suicide prevention development in China. Eight modules starting from improving suicide literacy to pragmatic techniques of gatekeeper behaviour were established, and delivery methods of the gatekeeper training program include videos, group discussion and role play.

Compared to other gatekeeper training programs, the Life Gatekeeper program featured a module where teachers would be trained to communicate with parents about their children’s suicidal risks. As mentioned previously, the Ministry of Education of China (2021) promoted school-family partnership in crisis intervention for students with suicidal risk, and emphasized that schools should assist parents to seek professional support for at-risk students promptly. This strategy has been supported by Chinese researchers, as the psychological crisis experienced by an adolescent potentially reflects a systemic crisis in their ecological system, and school teachers and parents play essential roles in protecting their safety and in supporting the recovery (Su, 2020). For example, the statement ‘*Teachers should communicate and update parents regularly about their children’s safety and what help the family may need*’ was endorsed by 97.1% of experts.

Considering that parents might have emotional reactions of fear, surprise, anger, helplessness and worries when being informed of their child’s suicidal risk by teachers (Ding, 2021), we included statements of ‘*When communicating with parents about their child’s suicide risk, teachers need to focus on parents’ emotions and inform them that suicide is largely preventable to prevent them from becoming overly anxious*’ and ‘*Teachers should talk to parents about the support available at school and the medical resources they can turn to in order to help alleviate excessive worry*’, and both statements were endorsed by 100% of panel members. Furthermore, since youth suicidality has been found to associate with family factors such as lack of parental warmth (Li, Wang & Bao, 2016), impaired family functioning (Leung, Kwok & Ling, 2016), perceived authoritarian parent and negative family climate (Lai & McBride-Chang, 2001), during the Life Gatekeeper training program, teachers will also practise explaining to parents the most appropriate approach to communicate with their at-risk child, such as the statements ‘*Teachers should remind parents to remain calm and listen patiently when their children reveal distressing emotions and thoughts and suggest they not blame, scold, or refuse to acknowledge their children’s suicidal thoughts or emotional distress*’ and ‘*Teachers should remind parents to take a cooperative rather than commanding approach when discussing solutions with their children about how they would like their families to support them*’, which both achieved 100% consensus among experts among first round of Delphi consensus.

Meanwhile, on the training version for parents, all statements achieved consensus by panel members. Statements in this section were about what and how parents should communicate with their children about their suicidal risks, and how they should seek further support from school, hospitals and local resources on the behalf of their children. Based on feedback from experts, a statement of life education was added in the second round, ‘*Parents need to educate their children about life and encourage them to discuss the value of life together*’, which was endorsed by 93.5% of panel members in the second round. Indeed, it was found that having meaning and purposes in life significantly impacted on attitude towards suicide among university students (Xie, Zou & Huang, 2012). Another statement stressing the importance of self-care of parents was added as proposed by experts and was endorsed by all panel members - ‘*Parents need to be in good shape to be competent in caring for their children. Thus, they also need to pay attention to their own emotions and learn to use internal and external resources to help themselves*’. As caregivers involved in suicidal crisis understandably are likely to feel distressed themselves, support for family members should also be promoted (Grant, Ballard & Olson-Madden, 2015). Nevertheless, this statement did not specify exact skill or technique that could help parents to manage their emotions, which could be elaborated into more details when developing the training for parents.

Half of the statements that did not reach consensus in the first round of Delphi were removed from the final section of ‘other reasons that prevent children from seeking help or teachers or parents from providing help’. For example, the statement ‘*The more severe the suicidal idea is, the more reluctant the individual is to seek help*’ which was only endorsed by 79.4% of experts, which described the help-negation effect pertinent to suicidal ideation (Wilson & Deane, 2010; Yakunina et al., 2010), and it was initially added with the intention to help gatekeepers understand it as a barrier that prevents individuals from seeking support. However, some experts expressed their concerns that this might not be accurate as many with severe suicidal ideation do seek help. Furthermore, the statement ‘*Fear of not being able to afford treatment is a realistic barrier that prevents individuals from seeking help*’ was only perceived as important/very important by 70.6% of panel members, yet there was no comment about this item in the feedback section. Finally, the statement of ‘*Having no time to go to the doctor is a realistic factor that prevents individuals from seeking help*’ was only endorsed by 61.8% of experts and were excluded at the first round, which reflected that a significant number of panel members did not consider it to be important for gatekeepers to learn about this barrier and no relevant feedback was given, despite that lack of time has been reported as a significant barrier to seek help by previous literature (Czyz et al., 2013; Hom, Stanley, & Joiner, 2015).

The rest of statements that did not reach consensus from the first round were risk factors related to suicide. For example, the statement ‘*Females have more suicidal ideation and suicide attempts than males, while males are more likely to commit fatal suicidal behaviors and die by suicide than females*’ was only endorsed by 73.5% of experts, although gender differences in suicidal behaviour have been consistently identified among Chinese adolescents (Liu et al., 2019; Zhang et al., 2019). It was commented by an expert that by emphasising more suicidal death by male than female student, it might cause teachers to underestimate the probability of suicide by female students. Meanwhile, for the other two statements ‘*Juvenile delinquency is a personal factor that increases suicide risk*’ and ‘*Excessive study pressure and anxiety before exams are school factors that increase the risk of suicide*’ which were only endorsed by 73.5% and 79.4% of panel members respectively, no specific comment was made in relation to them. However, there was feedback about the section that some items were more important for research but should not be specified in training, as these causes were too generic.

When developing statements for the current study, research literature from both English and Chinese database were consulted. As such, we could learn not only from evidence-based international gatekeeper programs (e.g. Holmes et al., 2021; Singer, Erbacher & Rosen, 2019), but also from Chinese literature that provided insights on socio-cultural factors relevant to the development of a localised gatekeeper training program in China, and from national guidelines of suicide prevention made by the government. For example, existing gatekeeper training programs such as ASSIST or QPR commonly require gatekeepers to refer at-risk students to school counsellor for support (Gould et al., 2013; Wyman et al., 2008). However, this might not be feasible in China due to the current limited availability of mental health professionals within schools, especially in rural areas (Zhao et al., 2017; Liang, Mays & Hwang, 2018). Therefore, we adapted the programme to this challenge by adding the school-parent partnership module and by providing a summary of crisis services and hospitals with mental health services available nationally and locally, to help teachers and parents feel confident in knowing where to seek support for at-risk students after attending the training.

As panel members were recruited from various provinces across the country, with diverse roles, education level and years of expertise in relation to suicide prevention, this added strength to the study as endorsed statements reflected consensus reached with comprehensive understanding of the topic, thus increased generalizability of the findings. Nevertheless, the acceptability of the Life Gatekeeper program by school teachers, parents and targeted adolescents has yet be assessed. The appropriate frequency to deliver this gatekeeper training remains unclear, and further investigation is warranted to evaluate the effectiveness of the Life Gatekeeper training program in promoting gatekeeper behaviour when being delivered to school teachers and parents.

This Delphi study provides a key foundation to promote the first localised and evidence-based suicide gatekeeper training program in Chinese schools for both teachers and parents. Although the content and delivery methods of this training share similarities with existing gatekeeper training programs, it also takes national guideline and sociocultural factors into consideration, and thus is more adaptive to be implemented in Chinese schools. In the future, the Life Gatekeeper training program could either be implemented by itself to reduce youth suicide in China, or it could also be used as part of a comprehensive school based suicide prevention program (Godoy Garraza et al., 2019; Goldston et al., 2010). We hope that the current study could pave the way for more evidence based and localised suicide prevention programs in China, which are much needed given the pressing concern of youth suicide within the country.

## Data Availability

All data produced in the present study are available upon reasonable request to the authors

## Conlifct of interst

None

## Data availability statement

Dr Runsen Chen had full access to all the data and takes responsibility for the integrity of the data

